# Allogenic Megakaryocyte Therapy in patients with Refractory Thrombocytopenia

**DOI:** 10.1101/2025.10.02.25336783

**Authors:** Aijie Huang, Liping Deng, Haiwei Liang, Bingbing Yan, Fei Wang, Guangyu Sun, Yan Zhang, Kaidi Song, Baolin Tang, Yongsheng Han, Xiang Wan, Wen Yao, Meijuan Tu, Ziwei Zhou, Yanxiao Ao, Jianan Zeng, Haochuan Gao, Jiali Zhao, Mingjie Yan, Kaini Liang, Yanan Du, Xiaoyu Zhu

## Abstract

**BACKGROUND:** Patients with refractory thrombocytopenia (RT) are not sensitive to conventional therapies, such as platelet transfusions and thrombopoietin receptor agonists (TPO-RAs). The persistently high risk of life-threatening hemorrhage in this population highlights the urgent need for novel therapeutic strategies. Megakaryocyte (MK)-based therapies have emerged as a promising alternative, as MKs are the natural precursor cells responsible for platelet production. However, whether allogeneic MK therapy can improve platelet counts and function in patients with RT remains unclear.

**METHODS:** We evaluated HLA-mismatched allogeneic MK therapy in 10 patients with RT following allogeneic hematopoietic stem-cell transplantation (allo-HSCT). All patients exhibited no response after at least one month of continuous treatment with TPO-RAs or other thrombopoiesis promoting therapies. MKs were expanded ex vivo and administered in a single infusion at one of three doses (1×10^6^, 5×10^6^, or 1×10^7^ MKs per kilogram of body weight). Safety and efficacy were closely monitored.

**RESULTS:** MK infusion had minimal impact on inflammatory cytokine levels and coagulation parameters of patients. Among the 10 patients treated, 8 (80%) demonstrated a clinical response; including 3 complete response, and 5 partial response. Clinical improvement was observed within 28 days after infusion across all dose levels.

**CONCLUSIONS:** Among 10 patients with RT, 8 responded to MK therapy without experiencing major toxic effects. (ClinicalTrials.gov number, NCT06534255.)

## INTRODUCTION

Refractory thrombocytopenia (RT) is a serious complication that may occur after intensive radiotherapy and chemotherapy[1] or allogeneic hematopoietic stem cell transplantation (allo-HSCT). In the post-transplant setting, RT, which is also referred to as refractory prolonged isolated thrombocytopenia (RPIT), is generally defined as a platelet count of <30×10^9^/L around day 60 post-transplantation, with no response after at least one month of treatment with recombinant human thrombopoietin (rhTPO), thrombopoietin receptor agonists (TPO-RAs), or other conventional measures such as glucocorticoids and intravenous immunoglobulin[2, 3]. The reported incidence of RT after allo-HSCT ranges from 20% to 37%, and this condition is strongly associated with transfusion dependence, increased morbidity, and a significantly higher risk of non-relapse mortality[4-6]. Due to persistently low platelet counts, patients with RT require constant platelet transfusions to reduce the risk of haemorrhage[7]. However, the effect of platelet transfusion is transient, and the repeated need for transfusion highlights the substantial unmet medical needs in this population.

In some cases, RT manifests as impaired platelet recovery after allo-HSCT, including delayed platelet engraftment (DPE) [8] or prolonged isolated thrombocytopenia (PIT) or secondary failure of platelet recovery (SFPR) [9]. Those complications are associated with inferior overall survival and transplantation-related mortality (TRM)[10-12]. Although TPO-RAs like Eltrombopag (EPAG), avatrombopag (APAG) and rhTPO have shown promise in some studies to promote platelet engraftment[10, 13, 14], these therapies rely on the patient’s own megakaryocytes (MKs) to exert the therapeutic effects. However, their clinical efficacy is highly heterogeneous among patients, and for those with low MK counts or functional abnormalities, achieving satisfactory outcomes remains challenging. Consequently, novel treatment strategies are urgently needed for this high-risk population.

Infusion of purified CD34+ cells has been shown to improve poor graft function (PGF) after allo-HSCT, indicating that the supplementation of hematopoietic progenitor cells may be a viable therapeutic approach[15]. While platelet transfusion remains the standard of care, their short duration of efficacy and limited supply underscore the need for alternative treatments. MKs, the bone marrow-resident cells, with the ability to give rise to platelet, may offer a novel therapeutic strategy. MKs have also been identified in the lungs and peripheral circulation[16]. In addition, single-cell RNA sequencing studies have revealed heterogeneity among MK subsets, suggesting functional diversity[17, 18]. Despite efforts to generate platelet in vitro using induced pluripotent stem cells (iPSCs) or CD34+ cells, progress remains limited by MK heterogeneity and the high production costs[19, 20]. The therapeutic potential of infusing megakaryocytic progenitor cells or mature MKs for RT remains largely unexplored. Therefore, we conducted a clinical study to evaluate the safety and efficacy of escalating doses of allogenic MKs in patients with RT.

## METHODS

### PATIENTS AND STUDY DESIGN

This report presents the initial findings from the MegaLT-2024 trial (NCT06534255), an open-label, single-arm, phase I study designed to evaluates the safety, efficacy, and pharmacokinetics of MegaLT in patients with RT following radiotherapy, chemotherapy, or allo-HSCT **(Fig. 1A)**. MegaLT is an off-the-shelf, cryopreserved allogeneic MK product formulated for clinical use. After obtaining written informed consent, ten patients with RT following allo-HSCT were enrolled and treated between December 2024 and May 2025. Two patients had undergone peripheral blood stem cell transplantation (PBSCT), while the remaining eight had received UCBT. All the patients met the inclusion criteria, with none meeting the exclusion criteria. At the data cutoff on July 31, 2025, the median follow-up duration was 191 days (range, 50–239 days). Briefly, patients were consecutively enrolled according to protocol, and received a single infusion of MegaLT at escalating doses of 1×10^6^, 5×10^6^, or 1×10^7^ MKs per kilogram of body weight **(Fig. 1B)**. Administered MegaLT products were partially human leukocyte antigen (HLA)-matched, with 1–5 of 10 matches at HLA loci A, B, C, DR and DQ) **(Table 1)**.

**Figure 1.**
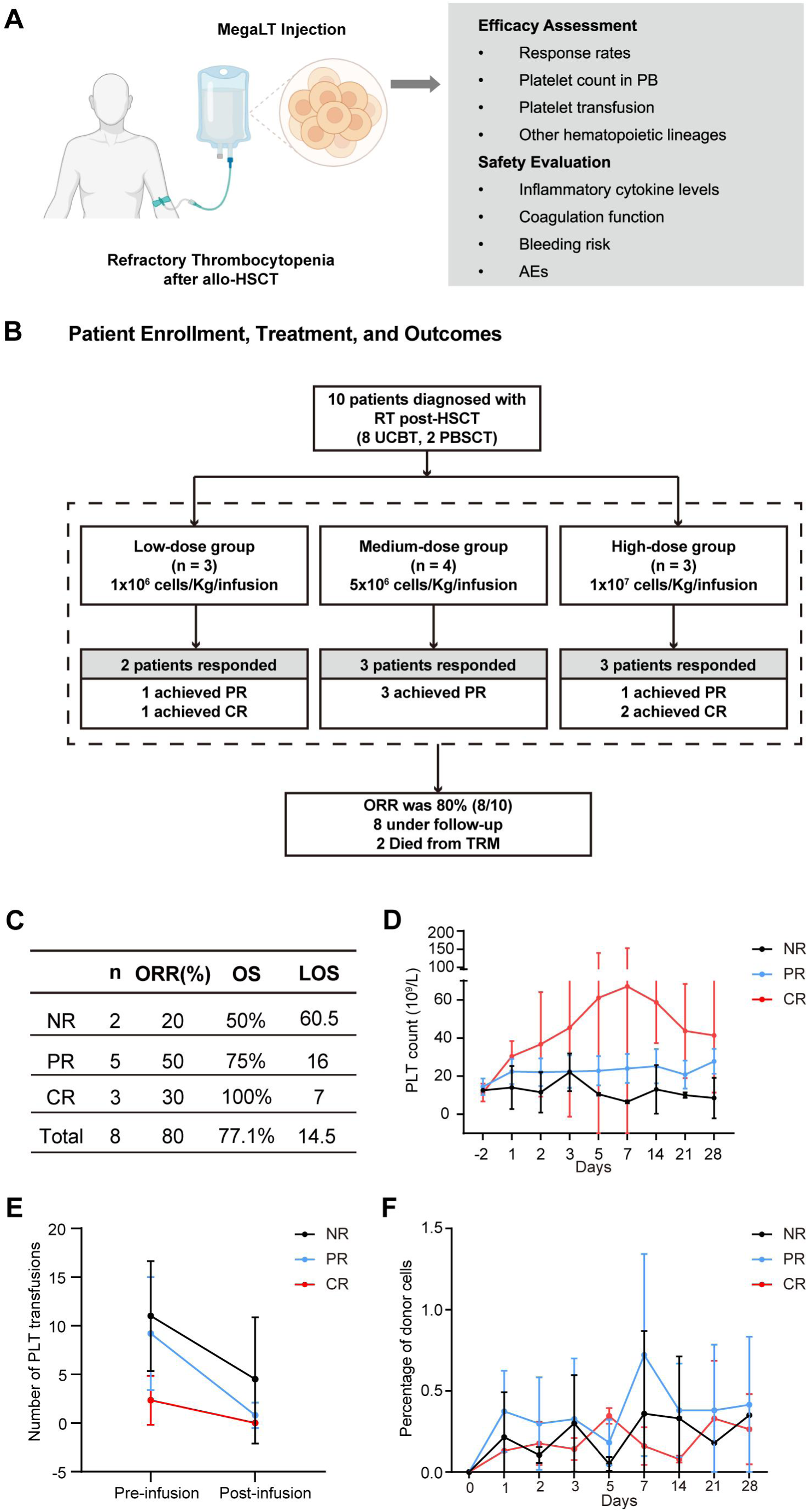
Clinical response of MegaLT. **A.** Overview of the treatment and evaluation strategy. The patient with refractory thrombocytopenia (RT) receives injection of the MegaLT (left). Efficacy and safety are evaluated using multiple assessment methods (right). **B.** Schematic illustration of patient enrollment, treatment, and outcomes. Ten patients with RT were enrolled and assigned to low-, medium-, and high-dose cell therapy groups. Partial and complete responses were observed across groups, yielding an overall response rate of 80%. Abbreviations: RT, refractory thrombocytopenia; HSCT, hematopoietic stem cell transplantation; UCBT, umbilical cord blood transplantation; PBSCT, peripheral blood stem cell transplantation; CR, complete response; PR, partial response; ORR, overall response rate; TRM, transplantation-related mortality. **C.** Table presenting patient numbers (n), overall response rate (ORR), overall survival (OS) rate, and length of stay (LOS). **D.** Platelet concentrations over time after MegaLT infusion. Black, blue, and red lines represent patients with no response (NR), partial response (PR), and complete response (CR), respectively. **E.** Platelet transfusion numbers pre- and post-infusion. Black, blue, and red lines indicate patients with NR, PR, and CR, respectively. **F.** Percentage of donor cells at different time points after infusion. Black, blue, and red lines represent patients with NR, PR, and CR, respectively.

**Table 1.** Baseline Characteristics and Clinical Outcomes.

| Patient No. | Dose<br>Level<br>*cells/kg* | Sex | Type of<br>Transpla<br>ntation | HLA Allelic<br>Match<br>*no./total no.* | Treatment<br>Efficacy |
| --- | --- | --- | --- | --- | --- |
| MegaLT01 | 1×10<sup>6</sup> | Male | UCBT | 2/10 | PR |
| MegaLT02 | 1×10<sup>6</sup> | Male | UCBT | 2/10 | CR |
| MegaLT03 | 1×10<sup>6</sup> | Male | UCBT | 1/10 | NR |
| MegaLT04 | 5×10<sup>6</sup> | Male | UCBT | 2/10 | PR |
| MegaLT05 | 5×10<sup>6</sup> | Male | UCBT | 1/10 | NR |
| MegaLT06 | 5×10<sup>6</sup> | Male | UCBT | 5/10 | PR |
| MegaLT07 | 5×10<sup>6</sup> | Male | UCBT | 4/10 | PR |
| MegaLT08 | 1×10<sup>7</sup> | Female | PBSCT | 1/10 | CR |
| MegaLT09 | 1×10<sup>7</sup> | Male | PBSCT | 2/10 | PR |
| MegaLT10 | 1×10<sup>7</sup> | Female | UCBT | 2/10 | CR |

*Abbreviations:* UCBT, umbilical cord blood transplantation; PBSCT, peripheral blood stem cell transplantation; CR, complete response; PR , partial response; NR, no response.

### STUDY OVERSIGHTS

Written informed consent was obtained from all participants prior to their inclusion in the study. The study was conducted in compliance with the principles of the Declaration of Helsinki and was approved by the Ethics Committee of the First Affiliated Hospital of the University of Science and Technology of China (Approval number, 2024KY-342).

### RESPONSE CRITERIA

A complete response (CR) was defined as a platelet count ≥ 50×10^9^/L following transfusion, with no requirement for platelet transfusion for at least seven consecutive days. A partial response (PR) was defined as an increase in platelet counts above baseline without reaching 50×10^9^/L, with no requirement for platelet transfusion for at least seven consecutive days. No response (NR) was defined as a platelet count below baseline at four weeks post-treatment or a persistent need for platelet transfusion. Overall survival (OS) was defined as the time from MegaLT infusion to death from any cause or last follow-up (July 31, 2025). Bleeding severity was graded using the modified World Health Organization (WHO) bleeding scale (Grades 0–4)[7]. Acute graft-versus-host disease (aGvHD) was diagnosed based on the Mount Sinai Acute GvHD International Consortium (MAGIC) criteria[21].

### SAFETY AND EFFICACY ENDPOINT

Safety assessments were performed at day 14 and day 28 post-infusion, with additional evaluations during long-term follow-up. Adverse events were graded according to CTCAE v5.0. The primary efficacy endpoint was the overall response rate (ORR), defined as the sum of patients achieving CR or PR, at week 4. Secondary endpoints included response rates at weeks 1, 2, 4, and 8; changes in platelet counts; time to the first platelet recovery (defined as platelet count ≥ 50×10^9^/L or ≥ 100×10^9^/L); achievement of transfusion independence within 4 weeks and recovery of other hematologic lineages (granulocytic and erythroid).

### STATISTICAL ANALYSIS

Overall survival was estimated using the Kaplan–Meier method. Additional statistical analyses are detailed in the figure legends. All analyses were conducted with the use of SPSS software (version 26) and GraphPad Prism (version 9.5).

## RESULTS

### CHARACTERISTICS OF THE PATIENTS

The cohort included seven patients with PIT after UCBT and three patients with SFPR, of whom two had undergone PBSCT and one had received UCBT. Underlying diseases consisted of acute myeloid leukemia (AML, n=4), myelodysplastic syndrome (MDS, n=3), acute lymphoblastic leukemia (ALL, n=1), plasma cell leukemia (PCL, n=1), and chronic myelomonocytic leukemia (CMML, n=1). The median age of the patients was 35 years (range, 10-69), and two were female. All patients were in complete remission at the time of study entry. At baseline, all patients exhibited severe thrombocytopenia requiring ongoing platelet transfusion. The median platelet count at enrollment was 12×10^9^/L (range, 6–20×10^9^/L), and bone marrow examination revealed markedly reduced or absent MKs in most patients. The median interval from transplantation to trial enrollment was 2.6 months (range, 2–45.7 months). No patients had active severe infections, uncontrolled GvHD, or evidence of significant organ dysfunction at the time of enrollment.

### EFFICACY

Following MegaLT infusion, 8 of 10 patients (80%) achieved an objective response, including 3 CR and 5 PR, while 2 patients showed NR. At the data cutoff, 8 patients were alive and remained under follow-up, while 2 had died due to transplantation-related mortality (TRM). As summarized in **Fig. 1C**, the ORR was 80% (8/10), with CR and PR rates of 30% (3/10) and 50% (5/10), respectively. Patients who achieved CR or PR demonstrated superior outcomes, with an overall survival (OS) of 100% and 75±21.7% respectively, compared with 50±35.4% in the NR group. The length of stay (LOS) followed a similar trend, with a median of 7 days in the CR group, 16 days in the PR group, and 60.5 days in the NR group **(Fig. 1C)**.

Peripheral blood platelet counts significantly improved in both CR and PR groups. Patients who achieved CR experienced a rapid increase in platelet counts, with peak levels observed around day 7 post-infusion, whereas PR patients showed moderate but sustained increases in platelet counts. In contrast, NR patients failed to achieve platelet recovery throughout the observation period **(Fig. 1D)**. Within 4 weeks of MegaLT infusion, three patients (MegaLT02, MegaLT08, and MegaLT10) achieved CR, with platelet counts reaching ≥ 50×10^9^/L, on days 13, 10, and 2, respectively. Notably, only MegaLT10 achieved a PLT count ≥ 100×10^9^/L, which occurred on day 5 **(Fig. S1A)**.

In addition to platelet recovery, improvements in other hematopoietic lineages were also observed **(Fig. S1B)**. Patients achieving CR demonstrated sustained increases in hemoglobin levels that remained consistently higher than those in the PR and NR groups throughout follow-up. Absolute neutrophil counts (ANC) gradually improved in both CR and PR patients but remained persistently low in NR patients. White blood cell (WBC) and lymphocyte (LYM) counts were relatively stable, with a tendency toward higher levels in CR patients compared with NR patients. Monocyte (MONO) counts fluctuated across all groups without significant differences. These findings suggest that MegaLT infusion contributed to multilineage hematopoietic recovery, particularly in patients who achieved CR.

During the 28 days prior to treatment, the median of platelet transfusions for all patients was 5.5 units (range, 0–16). Within 28 days after MegaLT infusion, the median number decreased to 0 units (range, 0–10). As shown in **Fig. 1E**, transfusion requirements were substantially reduced in patients who achieved CR or PR, while NR patients remained dependent on platelet transfusions. A similar reduction was observed in red blood cell transfusions **(Fig. S1C)**. These findings suggest that MegaLT infusion was associated with reduced transfusion burden among responding patients.

### PHRMACOKINETICS OF MEGALT

The pharmacokinetics of MegaLT were assessed using a digital droplet polymerase chain reaction (ddPCR) assay to detect donor-derived cells in peripheral blood post-infusion. The proportion of MegaLT donor cells ranged from 0.005% to 1.58% across all groups, with transient increases observed between day 3 and day 7 after infusion, particularly among PR patients. No sustained expansion of donor-derived cells was detected. Differences in cell persistence among CR, PR, and NR groups were modest, indicating no significant correlation between MegaLT persistence and the degree of HLA compatibility with the recipient **(Table 1 and Fig. 1F)**. Patients with a high degree of HLA compatibility (4–5/10 HLA loci) with MegaLT exhibited a higher proportion of detectable donor-derived cells, yet this does not correlate strongly with therapeutic efficacy **(Fig. S1D)**. In bone marrow samples, donor-derived cells exhibited a certain degree of chimerism (0.017%–0.64%). However, this level of chimerism did not correlate significantly with treatment efficacy **(Fig. S1E)**.

### PLATELET STRUCTURE AND FUNCTION

Following MegaLT infusion, elevated platelet levels were observed in responding patients. To investigate whether this increase was accompanied by improvements in platelet function, both platelet structure and function were evaluated before and after infusion. As the contents of platelet granules are essential for the hemostatic function of platelets, particular attention was given to the number of granules within each platelet. Transmission electron microscopy revealed that patients in the CR and PR groups demonstrated an increased number of platelet granules compared to those in the NR group, both before and after MegaLT infusion **(Fig. 2A)**. Prior to treatment, there were no statistically significant differences in granule count between groups. However, post-infusion analyses demonstrated a significant increase in granule count in the CR and PR groups compared to the NR group **(Fig. 2B)**, suggesting improved platelet functionality in responders. Furthermore, platelet structure appeared more intact across all groups following infusion. This was reflected by a reduced proportion of structurally abnormal platelets **(Fig. 2C)**, indicating enhanced platelet quality after MegaLT infusion. The improvement in platelet function following MegaLT infusion was furtherly substantiated by both flow cytometry **(Fig. S2A and S2B)** and thromboelastography analysis **(Fig. S2C)**.

**Figure 2.**
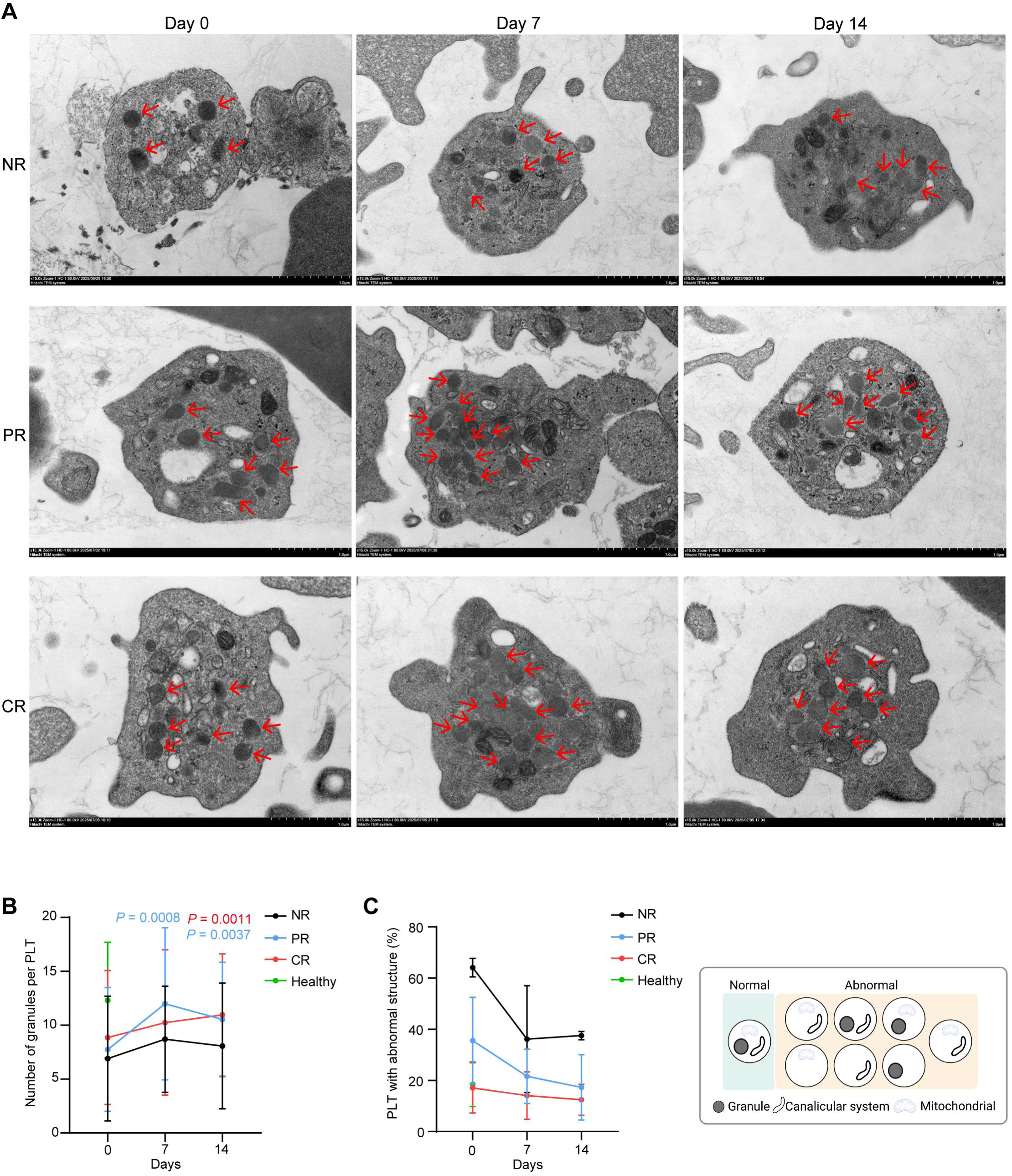
Evaluation of platelet structure and function in patients. **A.** Transmission electron microscopy (TEM) images of platelets from NR, PR, and CR patients at Day 0, Day 7, and Day 14 after treatment. The images clearly reveal platelet ultrastructures. Red arrows indicate granules of platelets. **B.** Number of granules per platelet at different time points after infusion. Black, blue, red, and green lines represent NR, PR, CR, and healthy controls, respectively. **C.** Percentage of platelet with abnormal structure at different time points after infusion. Black, blue, red, and green lines represent NR, PR, CR, and healthy controls, respectively. The diagram on the right illustrates the differences between normal and abnormal platelet structures.

### SAFETY PROFILES

Following MegaLT infusion, none of the patients exhibited symptoms consistent with cytokine release syndrome **(Fig. 3A).** Serial monitoring of coagulation parameters revealed that the MegaLT infusion had no significant impact on coagulation function in responders (CR and PR). Prothrombin time (PT), activated partial thromboplastin time (APTT), thrombin time (TT), and fibrinogen (Fg) levels remained stable throughout follow-up, with no indication of treatment-related coagulopathy **(Fig. 3B)**. In contrast, one non-responder (MegaLT05) exhibited prolonged PT and APTT, suggesting impaired coagulation function and increased bleeding risk. Collectively, these findings indicate that MegaLT infusion was not associated with coagulation abnormalities in responders, whereas coagulation dysfunction in non-responders more likely reflected disease persistence rather than treatment toxicity.

**Figure 3:**
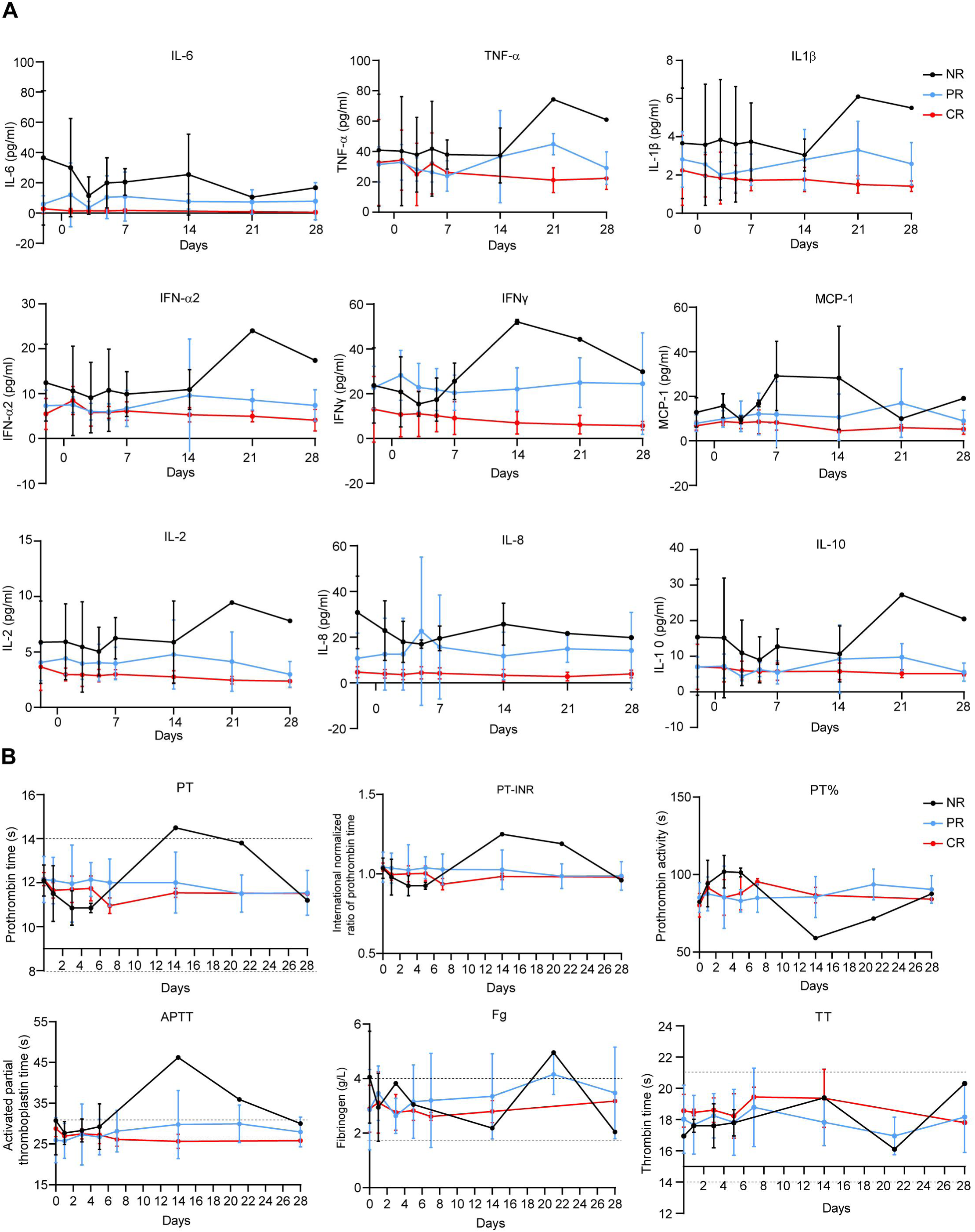
Safety profile of MegaLT therapy in patients. **A.** Changes in serum cytokine levels before and after treatment. Line plots show concentrations of IL-6, TNF-α, IL-1β, IFN-α2, IFN-γ, MCP-1, IL-2, IL-8, and IL-10 measured in serum at the indicated time points. Black, blue, and red lines represent NR, PR, and CR, respectively. **B.** Dynamic changes in coagulation-related factors, including prothrombin time (PT), prothrombin time–international normalized ratio (PT-INR), prothrombin activity (PT%), activated partial thromboplastin time (APTT), fibrinogen (Fg), thrombin time (TT). Black, blue, and red lines represent NR, PR, and CR, respectively. Dashed lines denote the normal reference ranges for each parameter.

Bleeding risk, as assessed by the World Health Organization (WHO) bleeding scale, showed improvement following treatment **(Table 2)**. At baseline, five patients presented with Grade 1 bleeding. After infusion, bleeding risk improved to Grade 0 in two of these patients (one CR and one PR) within 14 days after infusion. Among non-responders, MegaLT05 remained at Grade 1, whereas MegaLT03 progressed to Grade 3 due to gastrointestinal bleeding in the third week post-infusion. One other patient, who achieved PR, remained at Grade 1 throughout the first 28 days post-infusion. Overall, bleeding progression to Grade 3 was only observed in the NR group. These results suggest that MegaLT infusion was associated with a reduced bleeding risk in responders, consistent with observed improvements in platelet counts and decreased transfusion dependence.

**Table 2.**
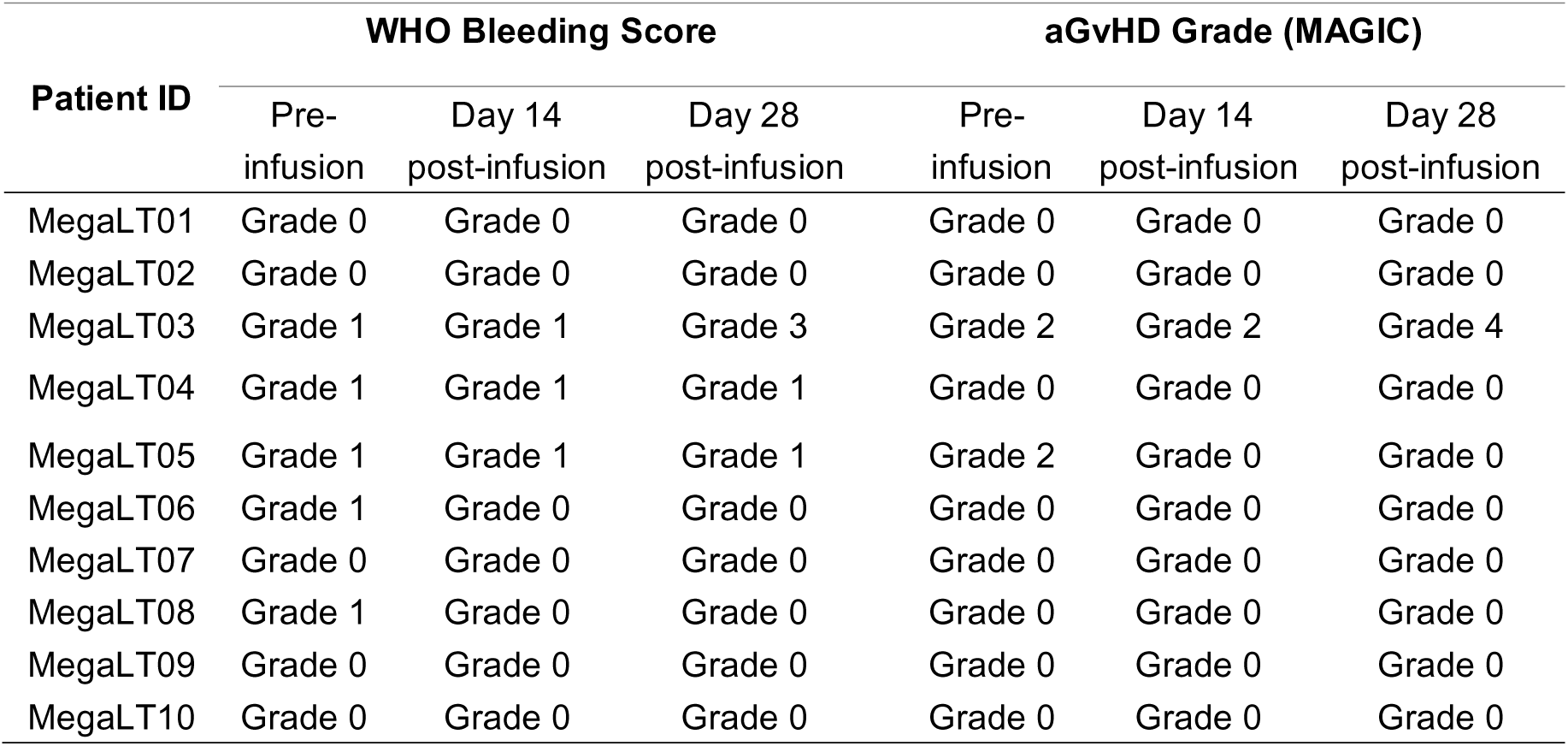
Safety Outcomes: Bleeding Score and aGvHD Status.

Among the ten treated patients, two experienced changes in aGvHD grade after infusion **(Table 2)**. MegaLT03 developed gastrointestinal bleeding, which was classified as stage IV aGVHD based on the MAGIC criteria. However, these events were considered unrelated to the MegaLT infusion. In contrast, MegaLT05 demonstrated an improvement in aGVHD status, decreasing from grade 2 to grade 0 post-infusion. Overall, MegaLT treatment was generally well tolerated **(Table 3)**. The most commonly reported adverse events (AEs) within 28 days post-infusion were pyrexia (3/10, 30%), abdominal pain (2/10, 20%), and diarrhea (2/10, 20%). Gastrointestinal hemorrhage occurred in one patient (10%). Most AEs were classified as mild to moderate (Grade 1–2). One patient, MegaLT03, experienced Grade 3 gastrointestinal hemorrhage, which was deemed unrelated to the MegaLT injection. Importantly, no treatment-related serious AEs or dose-limiting toxicities (DLTs) were observed during the study.

**Table 3.**
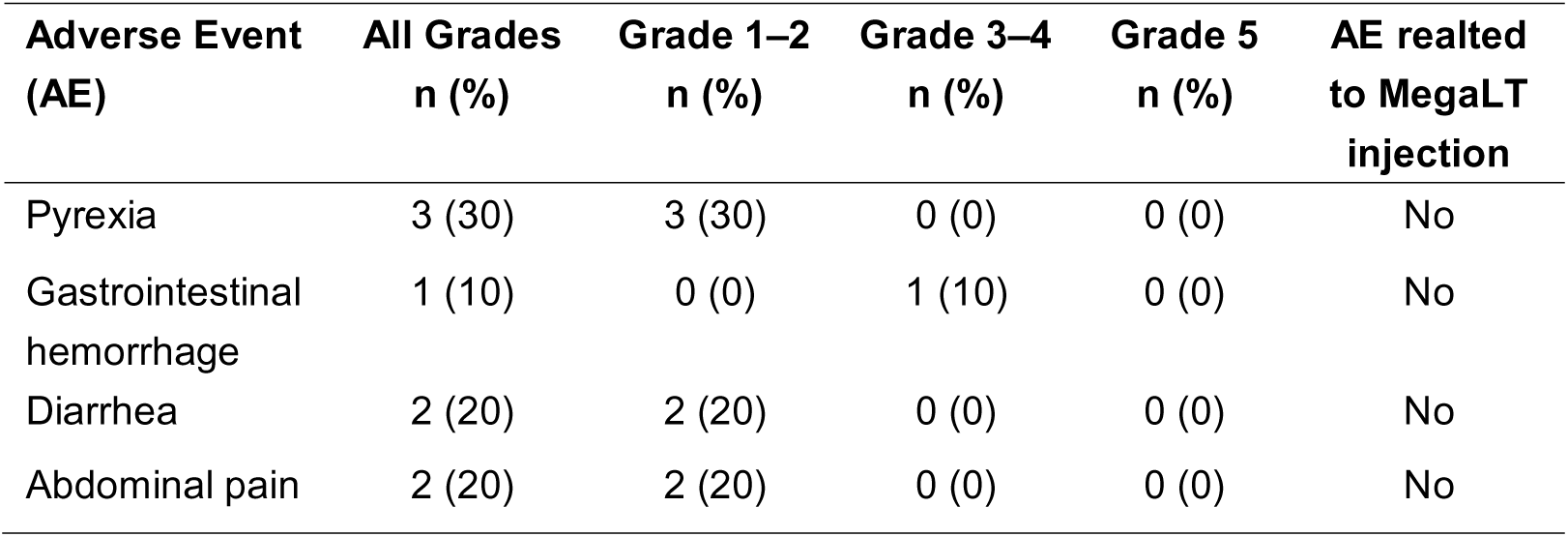
Summary of Adverse Events in the Study Population Within 28 Days Post-Infusion (Graded per CTCAE v5.0).

Two deaths occurred among the enrolled patients. MegaLT03 died in the second month post-infusion due to severe pulmonary infection and respiratory failure, whereas patient MegaLT04 died approximately three months post-infusion due to relapse of the primary disease.

## DISCUSSION

We report early clinical outcomes of allogenic MK therapy derived from cord blood in treating patients with RT following allo-HSCT. At a median follow-up of 191 days, 8 of the 10 treated patients (80%) achieved an objective response, and 3 of the 10 patients (30%) had a complete response. The treatment was generally safe and well-tolerated. The primary adverse events were fever and diarrhea, which were determined to be unrelated to MK infusion.

Patients enrolled in this study exhibited platelet counts as low as 30×10^9^/L or were dependent on platelet transfusions. Multiple platelet transfusions[22] and extended treatment with TPO-RAs, or other platelet-elevating therapies over one month were ineffective. In contrast, MegaLT therapy achieved an ORR of 80%, representing a promising treatment option for patients with RT. While EPAG has shown efficacy in post-transplant PGF and in the trilineage hematopoiesis in refractory severe aplastic anemia[23, 24], its clinical benefit in RT has been limited. A second infusion of CD34+ cells from the original donor may be considered for RT; however, this approach entails a prolonged differentiation process[25, 26], making it unsuitable for rapid platelet recovery. Furthermore, the isolation and purification of CD34+ cells using GMP-grade magnetic beads is both technically demanding and costly.

Within the bone marrow microenvironment, MKs play a crucial role in maintaining both structural and functional integrity[27, 28]. It has been demonstrated that MKs help preserve hematopoietic stem cell (HSC) quiescence and promote their regeneration following injury by secreting factors such as CXCL4 (PF4) and TGF-β [29, 30]. Our results suggest that higher MK counts in the bone marrow may be associated with improved treatment outcomes. Similarly, patients who presented with higher baseline haemoglobin levels tended to have better therapeutic responses, offering a potential metric for future patient selection.

MegaLT pharmacokinetics demonstrated that donor MKs did not undergo extensive proliferation post-infusion and maintained a stable proportion throughout the observation period. This suggests that low HLA compatibility does not result in rapid rejection of the infused allogenic cells. Pharmacokinetic profiles were comparable among patients with differing therapeutic responses, suggesting that individual variation in response to MK therapy likely contributes to clinical outcomes. Patients in all dose cohorts responded, and no treatment-limiting toxicity was observed, even in the high-dose group. Notably, allogeneic MKs did not induce cytokine release syndrome, a complication frequently associated with CAR T-cell therapy [31, 32]. Adverse events were mostly mild to moderate in severity (Grade 1–2). No treatment-related serious AEs or DLTs were reported. However, one patient experienced an increase in bleeding scores post-infusion, attributed to transplant-related complications rather than treatment toxicity. A predefined strategy for AE management was in place, although it was not required during this study.

Our findings suggest that MK therapy may serve as a preferred therapeutic option for RT, potentially redefining the therapeutic paradigm. Patients with RT demonstrated a response rate as high as 80% to MK therapy. Furthermore, its application may be extended to patients with non-refractory thrombocytopenia, where hematopoietic function is relatively preserved and therapeutic efficacy could be even greater. Unlike platelet transfusions or TPO-RAs, which offer only transient or limited benefits, MK therapy may exert dual action through direct platelet supplementation and regulation of the bone marrow microenvironment[33]. Future directions include the integration of MK infusion with agents such as TPO-RAs to enhance clinical outcomes.

While the initial findings are encouraging, limitations include the small sample size and limited follow-up duration. Larger-scale investigations, including randomized controlled trials, are necessary to further validate the efficacy and safety of this novel therapy. Furthermore, the molecular mechanisms underlying the observed clinical benefits remain incompletely understood, underscoring the need for additional preclinical research. Beyond RT, the potential therapeutic scope for MK therapy includes adjunctive use in HSCT and treatment of aplastic anemia or immune thrombocytopenia[34-36], for which more effective options are still needed. In conclusion, our study demonstrates that allogenic MK therapy yields substantial clinical benefits and has a favorable safety profile in RT patients. These findings support the ongoing development and a larger clinical evaluation of this promising therapeutic modality.

## Data Availability Statement

Due to patient privacy and ethical restrictions, the clinical datasets are not publicly available. De-identified data may be obtained from the corresponding author on reasonable request and with approval from the Ethics Committee.

## Figure legends

**Supplementary Figure 1.**
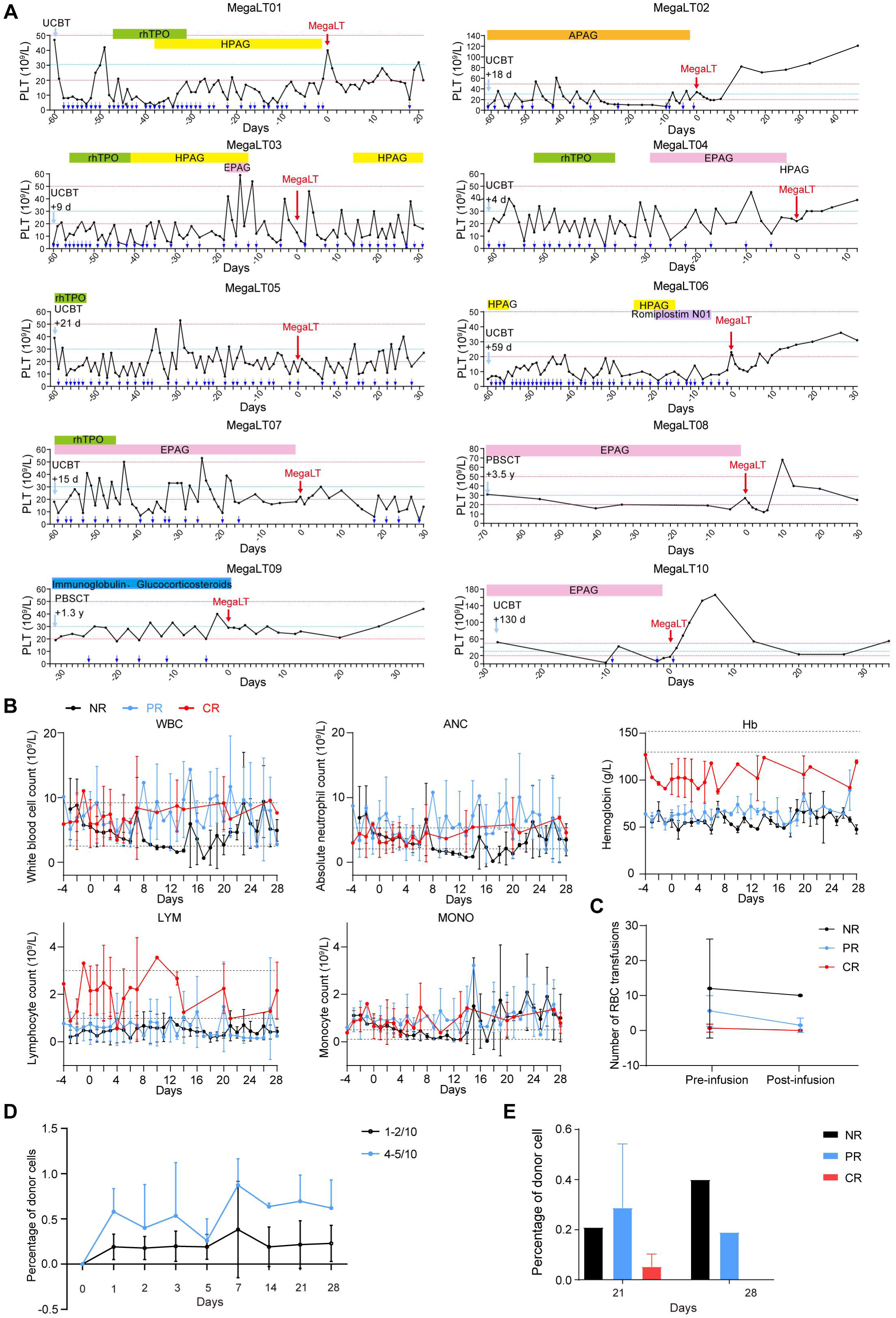
Clinical response of MegaLT. **A.** Peripheral blood platelet counts in 10 patients before and after MegaLT injection. The x-axis shows time relative to treatment, with Day 0 marking the MegaLT injection (indicated by a red arrow). The y-axis indicates platelet counts. Blue arrows denote platelet transfusion events, and horizontal-colored bars represent additional treatments administered during the study period. The red dashed lines indicate platelet levels of 20 and 50 (×10^9^/L), and the blue dashed line indicates a platelet level of 30 (×10^9^/L). **B.** Dynamic changes of peripheral blood parameters, including white blood cell count (WBC), absolute neutrophil count (ANC), lymphocyte count (LYM), monocyte count (MONO), and hemoglobin (Hb) throughout MegaLT therapy. Black, blue, and red lines represent no response (NR, n = 2), partial response (PR, n = 5), and complete response (CR, n = 3), respectively. Dashed lines denote the normal reference ranges for each parameter. **C.** Red blood cell (RBC) transfusion numbers pre- and post-infusion. Black, blue, and red lines indicate patients with NR, PR, and CR, respectively. **D.** Percentage of donor cells at different time points after infusion. Black and blue lines indicate patients with different HLA compatibility with MegaLT. **E.** Percentage of donor cells in bone marrow at Day 14 and Day 28 after infusion. Black bars represent NR, blue bars represent PR, and red bars represent CR.

**Supplementary Figure 2.**
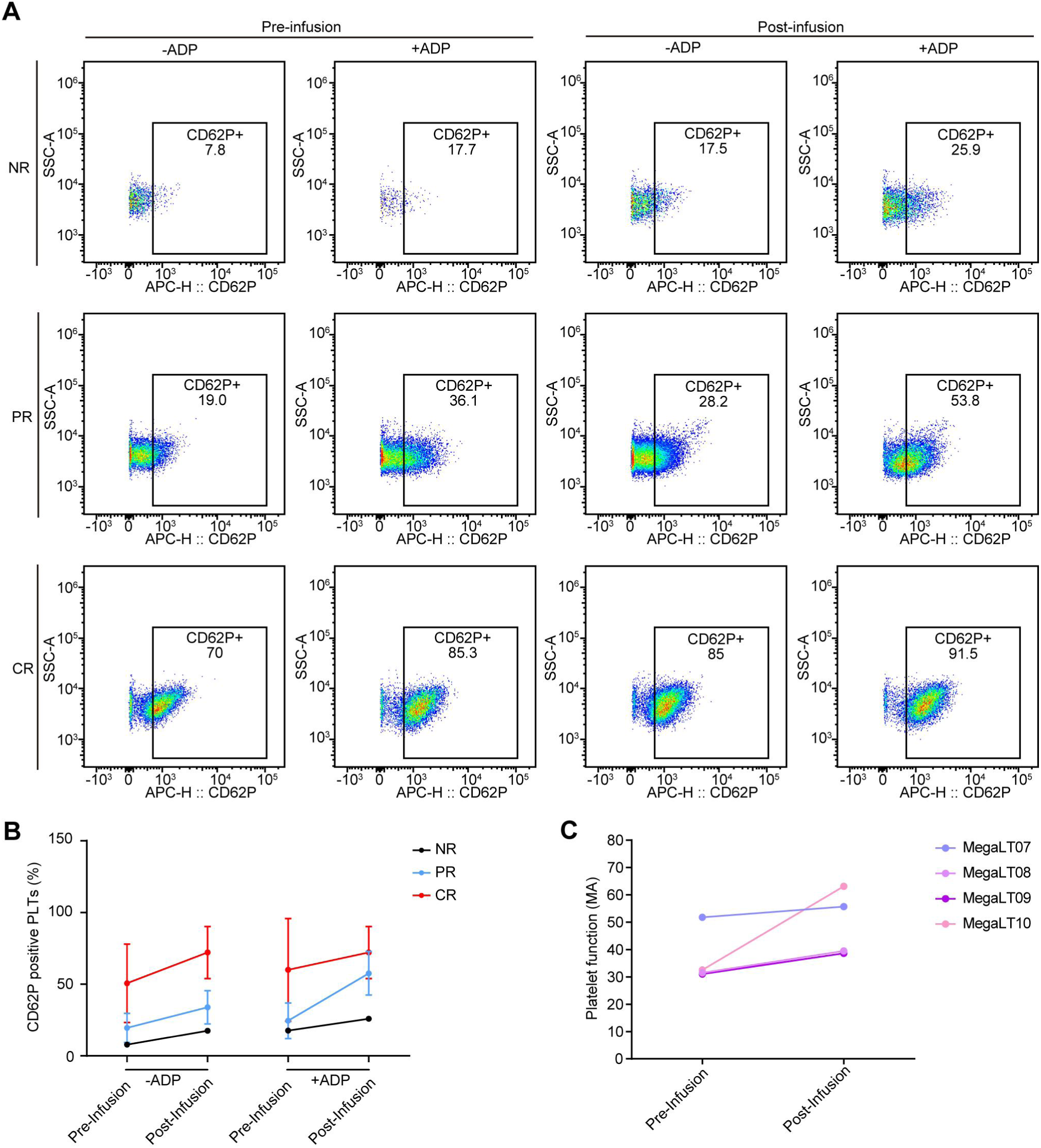
Evaluation of platelet function in patients. **A.** Flowcytometry detection of platelet activation ability pre- and post-infusion of NR, PR, CR, respectively. The flow cytometry results shown representative outcomes of platelet activation ability for each group. **B.** Statistical data for CD62P-positive platelets in the NR, PR, and CR groups with or without ADP. **C.** Thromboelastogram results of patients pre- and post-infusion. Platelet function has been improved after MegaLT infusion.

